# Redefining and estimating the early-phase reproduction ratio for epidemic outbreaks in spatially structured populations

**DOI:** 10.64898/2026.01.26.26344841

**Authors:** Boxuan Wang, Eugenio Valdano

## Abstract

Assessing epidemic risk following pathogen introduction is crucial in infectious disease epidemiology. Risk is commonly encoded through reproduction ratios, which underpin operational decision-making. In spatially structured populations, both local and cross-community transmission shape epidemic trends, a feature that standard reproduction ratios fail to capture simultaneously. Here, we use multitype branching processes to define the outbreak reproduction ratio *R*^ob^, a reformulation applicable across pathogens, epidemics and transmission routes, enabling community-specific, but system-aware, risk assessment. We test *R*^ob^ on respiratory pathogens and estimate it prior to emergence using aggregated contact matrices, enabling spatially resolved risk assessment even with limited data and computational resources. Estimates across countries reveal heterogeneous spatial risk, not captured by standard metrics. *R*^ob^ can also be estimated from early-phase surveillance data, as we show using SARS-CoV-2 in Canada, where it correctly identifies community risk. *R*^ob^ represents a concise and practicable framework for interpreting epidemic risk in spatially structured populations.

## Introduction

Assessing and predicting the conditions that determine whether a newly emerged or introduced pathogen will trigger a large-scale epidemic is central to epidemic preparedness [1]. One of the most popular metrics in epidemiology to assess epidemic risk is the reproduction ratio, *R*, which encodes the number of secondary infections in transmission chains [2]. Importantly, if *R* is above one, the outbreak may grow into a large-scale epidemic, and the probability of that happening, and the size of the resulting outbreak, increase with the value of *R* [3]. The reproduction ratio has been used across epidemics, diseases and regions to both understand and predict the effect of public health interventions. Examples abound. The very high reproduction ratio of measles has long driven vaccination policies targeting ≥ 95% coverage [4]. During the 2014-2016 Ebola outbreak in West Africa, estimates of *R* were crucial to assess population-level interventions [5]. In 2020, early estimates of the reproduction ratio of COVID-19 long-range and short-range mobility restrictions across the globe [6–8]. Recently, *R* is increasingly being used to monitor arboviral risk (dengue, chikungunya, Zika) driven by climate change [9].

Existing theoretical frameworks broadly estimate the reproduction ratio either from statistical inference on surveillance data, e.g., timeseries of reported cases [10–15], from transmission models that integrate contact and mixing data of the population [16–21], or more recently on phylogenetic data [22].

Estimating the reproduction ratio before, or at the start of, a new outbreak is particularly important to gauge its potential to cause a large-scale epidemic. It is also when it is the hardest to estimate, because this is when epidemic evolution is most sensitive to stochastic effects [23, 24] and heterogeneities in the underlying contact structure [25–27].

The challenge becomes even greater in spatially structured populations, composed of distinct communities connected by mobility. Existing frameworks face two main limitations. First, those providing local estimates of *R* neglect that infections arise not only from local transmission but also from mobility-driven interactions between communities [9, 28]. When this distinction is made, models typically assume that transmission is already established at the system level [29], or they separate only local and imported cases [30–33], without capturing the source-sink dynamics sustaining epidemic emergence and persistence [34, 35] or the long-range spread to previously unaffected regions [36, 37]. Second, frameworks that explicitly incorporate spatial, mobility-driven contact networks typically yield a single system-wide value of *R* [15, 17, 38]. Such global metrics provide limited information on the epidemic outcome following introduction in a specific community, failing to identify potential outbreak hotspots and offering only a global risk measure once circulation is already established. As a result, obtaining local yet system-aware estimates of epidemic risk for emerging outbreaks often depends on large computational models that require extensive, heterogeneous data on disease natural history, host behavior, and mobility [39, 40]. Such data are frequently unavailable, time-consuming to collect, and computationally expensive to process, delaying or preventing timely assessment in resource-limited settings. Moreover, the resulting metrics may often be less generalizable and harder to interpret than the reproduction ratio, limiting their practical use by public health authorities.

It is therefore essential to develop a theoretical framework that jointly captures local transmission conditions and the meso- and large-scale structures linking communities through mobility. To this end, we introduce the outbreak reproduction ratio (*R*^ob^): a parsimonious, interpretable, and computationally efficient indicator for rapid assessment of epidemic potential. *R*^ob^ quantifies outbreak risk as a function of the site of pathogen emergence or introduction, incorporating both local dynamics and system-level source-sink effects, and can be estimated either preemptively—before the pathogen arrives, or during the early stages of an outbreak.

We define *R*^ob^ using multi-type branching processes and demonstrate that it can be estimated both from low-resolution, aggregated spatial contact data collected pre-emptively before the outbreak, as well as from early epidemic surveillance data.

We validate the newly defined *R*^ob^ through synthetic epidemics informed with data-driven spatial contact networks, and we apply it to three case studies to showcase its potential. First, we simulate an epidemic of a directly transmitted respiratory pathogen in Italy and show that *R*^ob^ can be accurately estimated from surveillance data, and that it reveals systematic biases in traditional epidemiological indicators. Second, we extend the analysis across multiple countries again using data-driven contact networks, linking *R*^ob^ to demographic and spatial patterns and highlighting the limitations of both local and global reproduction metrics in structured populations. Finally, we study early SARS-CoV-2 transmission in Canada using transmission chain data and demonstrate that *R*^ob^ correctly identifies outbreak hotspots consistent with historical epidemic outcomes.

## Results

To model the early evolution of epidemic outbreaks, we use branching processes – a standard framework that explicitly captures the stochastic dynamics dominating the initial phase of epidemic spread [25]. We consider a population composed of *N* spatial communities, where transmission can occur locally in each community and also across communities. We encode spatial transmission patterns in the *N* -dimensional *reproduction operator* **R**, whose entry *R*_*ij*_ encodes the expected number of secondary infections that an infected resident of *j* generates among residents of *i* [15]. The spectral radius of **R**, which under realistic conditions is also a positive eigenvalue, is the reproduction ratio of the system, which, following Ref. [15], we will call *reference reproduction ratio R*^ref^, and determines the dynamics of the epidemic once it is established.

Notably, the reproduction operator is independent and agnostic of the diseases’s natural history and the demographic structure of the population. It can thus be applied to any transmissible disease and transmission route —-direct, vector-borne, or otherwise – provided that sufficient data exist to estimate its entries reliably. In the case studies below, we show how **R** can be reconstructed and used for directly transmitted respiratory pathogens.

We can formalize infection occurring within and across spatial communities as a multitype branching process [41], where the *type* of the offspring encodes the spatial communities in which the infection is generated.

To quantify the epidemic potential of a pathogen emergence (or introduction) event in community *i* we wish to compute the probability *p*_*i*_ that this leads to a major epidemic in the system (*epidemic probability* ). This is, in our framework, the probability that the branching process does not go extinct. We remark that the branching process is a good model of disease spread only before **R** effectively changes due to the epidemic dynamics (e.g., the accumulation of immunity) or to external interventions. *p*_*i*_ thus a good indicator for the early evolution, which is of interest of us, telling us whether the outbreak will reach the epidemic phase or not.

Assuming that infections generated in different communities by the same index case are independent, and the number of secondary infections depends only on its expectation value *R*_*ji*_ plus, possibly, other parameters that are constant across the system, the epidemic probability obeys the following equation:

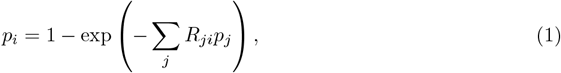

Derivation of this equation is shown in the Methods. We assumed that the offspring distribution is Poisson. This choice is reasonable despite the well-documented overdispersion of secondary infections in many epidemics, because overdispersion is often introduced as an effective proxy for effects due to structured populations, which we already model explicitly [22, 25]. These structures can be purely spatial, which we explicitly consider in this study, or arise from other sources such as household or classroom clustering [42]. Our framework is agnostic to the origin of the underlying structure and can also be applied to alternative forms of heterogeneity [20]. As a result, most additional sources of overdispersion can be incorporated through the explicit modeling of the relevant contact structure, without requiring changes to the theoretical formulation.

In the case of no inter-community transmission (*R*_*ij*_ = 0 ∀*i* ≠ *j*), the epidemic probability is well-known [3], and has a closed-form analytical expression: 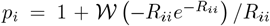, where 𝒲 is the principal branch of the Lambert-W function. Specifically, when *R*_*ii*_ *<* 1, the epidemic probability is zero, as no major epidemic can occur below the epidemic threshold. Above *R*_*ii*_ = 1, the epidemic probability increases sharply, and monotonously, with *R*_*ii*_. For this reason, in isolated communities the local reproduction ratio *R*_*ii*_ is also a reliable indicator of an outbreak’s potential to develop into a major epidemic, and is routinely used for this purpose [9]. Let us now assume that infected residents of community *i* can also transmit infection to residents of another community *j* (*R*_*ji*_ *>* 0, *R*_*jj*_ *> R*_*ii*_), and that no other inter-community connections exist –this is effectively a simplified two-community toy model where residents of *i* can generate infections in *j* and not viceversa. In this case, the epidemic probability in *i* is 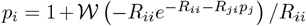. Figure 1a shows *p*_*i*_ as a function of the coupling term *R*_*ji*_. It becomes positive even when *R*_*ii*_ *<* 1, indicating that the local reproduction ratio is no longer a reliable measure of epidemic risk. At the same time, *p*_*i*_ differs from *p*_*j*_ and remains small across a wide range of *R*_*ji*_ values, showing that the system’s reference reproduction ratio – equal to *R*_*jj*_ in this simple case – is also an unreliable predictor of outbreak probability in *i*. Finally, Figure 1a illustrates that the total average number of secondary infections generated by residents of *i* (*R*_*ii*_ + *R*_*ji*_) likewise fails to capture epidemic risk.

**Figure 1.**
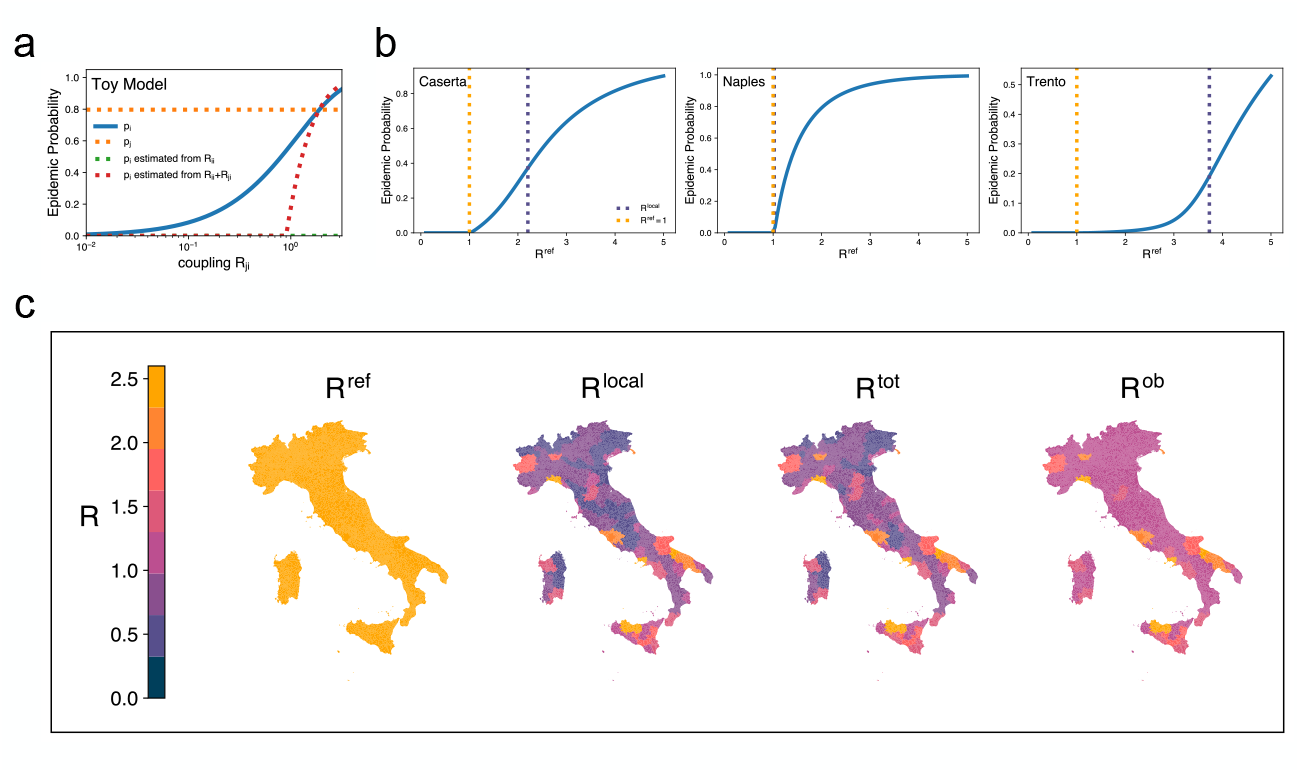
Epidemic probability and outbreak reproduction ratio *R*^ob^. **a**. Epidemic probability in community *i* (*p*_*i*_) in a two-community toy model where *i* is connected to *j* with coupling strength *R*_*ji*_: average number of infections that an infected resident of *i* generates in *j*. The x-axis is *R*_*ji*_ and the other parameters are fixed to *R*_*ii*_ = 0.1 and *R*_*jj*_ = 2.**b**. Epidemic probability in the Italian provinces (ADM-2) of Caserta, Naples, Trento, computed using colocation data and plotted against the country’s system-level reference reproduction ratio (*R*^ref^). Dashed lines indicate *R*^*ref*^ = 1 (global epidemic threshold) and local threshold values (*R*_*ii*_ = 1). **c**. Reproduction ratio estimates in Italian provinces assuming *R*^*ref*^ = 2.5. Different maps display different indicators and notably compare the outbreak reproduction ratio *R*^*ob*^ to commonly used risk metrics: *R*^ref^, the local *R* defined as the average number of locally generated infections, and *R*^tot^ defined as the average number of infections generared anywhere by a resident of a specific community (see main text for mathematical definitions).

These observations extend to complex spatial structures. In Figure 1b, we estimated the epidemic probability of a directly transmitted respiratory pathogen (such as influenza, SARS-CoV-2, or a newly emerging strain) across Italian provinces (ADM-2 level) for varying reference reproduction ratios. We used spatial contact data provided by Meta to model the reproduction operator **R**. Details are provided in Methods. Figure 1b shows that epidemic risk varied markedly across communities, and that neither local (*R*_*ii*_) nor global (*R*^ref^) estimates reliably capture it. In the province of Trento, the epidemic probability remains near zero even when the system reproduction ratio *R*^ref^ is well above one, yet rises significantly before the local *R*_*ii*_ exceeds unity, showing that both global and local measures may misrepresent risk. Naples —-one of Italy’s largest cities – exhibits the opposite pattern, with the epidemic probability increasing sharply as soon as *R*^ref^ *>* 1. Caserta, adjacent to Naples, shows intermediate behavior: *p*_*i*_ becomes positive just above *R*^ref^ = 1 but grows more slowly than in Naples, and without a clear relation to *R*_*ii*_. Together, these results demonstrate that both local and system-level reproduction ratios can yield misleading assessments of epidemic risk in spatially structured populations.

### The outbreak reproduction ratio *R*^**ob**^

These findings show that the definitions of the reproduction ratio based trivially on the average numbers of secondary infections – namely the local reproduction ratio *R*_*ii*_ and the total reproduction ratio 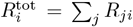 – fail to capture the contribution of mobility-driven transmission. Conversely, system-level measures such as *R*^ref^ overlook local heterogeneities that critically shape epidemic risk.

To overcome this, we introduce the *outbreak reproduction ratio* (*R*^ob^) as the average difference between the number of secondary infections needed to set off a large-scale epidemic and the number of secondary infections that lead to a minor outbreak. Consider an index infection in community *I* and denote its number of secondary infections in *j* as *X*_*i*→*j*_. By definition, *R*_*ji*_ = 𝔼 [*X*_*i*→*j*_]. Then, 𝔼 [*X*_*i*→*j*_|epidemic] and 𝔼 [*X*_*i*→*j*_|extinction] as the expected values conditioned on the two epidemic outcomes. In the framework of the branching process the distinction is on whether the process goes extinct or not, the latter happening with probability *p*_*i*_. In the case of realistic epidemic process we will show that this distinction can be made by choosing a cutoff on the attack rate and the empirical means are weakly sensitive to the chosen cutoff. The formal definition of the new metric is thus

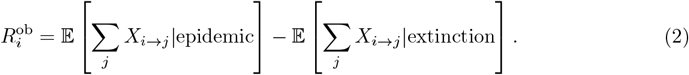

To study the properties of the outbreak reproduction ratio it is convenient to prove two equivalent definitions in terms of epidemic probabilities (proof in Methods):

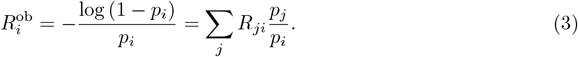

The first expression in Eq. (3) shows that the outbreak reproduction ratio corresponds to the local reproduction ratio that community *i* would require, if isolated, to yield the same epidemic probability. *R*^ob^ therefore represents an enhanced form of the local reproduction ratio *R*_*ii*_, incorporating nonlocal transmission effects into a single, community-specific metric. The second expression in Eq. (3) shows that *R*^ob^ equals the sum of the average number of infections generated by residents of *i* across all communities, each weighted by the relative outbreak risk of those communities with respect to *i*. In this sense, *R*^ob^ generalizes the total reproduction ratio 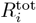, weighting infections according to their potential to initiate a large-scale epidemic rather than treating all secondary transmissions equally. For example, generating infections in a community (*j*) where the probability of onward epidemic is substantially larger than the local probability (*p*_*j*_ ≫ *p*_*i*_) boosts the outbreak reproduction ratio of *i*, showing that local estimates *R*_*ii*_ may substantially underestimate epidemic risk. At the same time, if the probability of outbreak in *j* is small (*p*_*j*_ ≪ *p*_*i*_) it will contribute marginally to risk in *i* (i.e., to 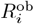), even if *R*_*ji*_ is large and contributes substantially to 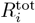.

We validated Eq. (3) against the simulated early epidemic phase of a directly-transmitted, respiratory pathogen spreading in Italy, using the same data as in Fig. 1 see details in the Methods. Importantly, we set a cutoff *M* on the total size of the epidemic to discriminate between epidemics (size exceeding *M* ) and minor events (size below *M* ). We computed *R*^ob^ from Eq. (2) using sample means from different stochastic realizations, then compared it to the one obtained via the epidemic probabilities in Eq. (3). Fig. 2a shows the comparison for six Italian provinces, proving that they match for cutoff values spanning orders of magnitudes, and even for relatively low cutoff values.

**Figure 2.**
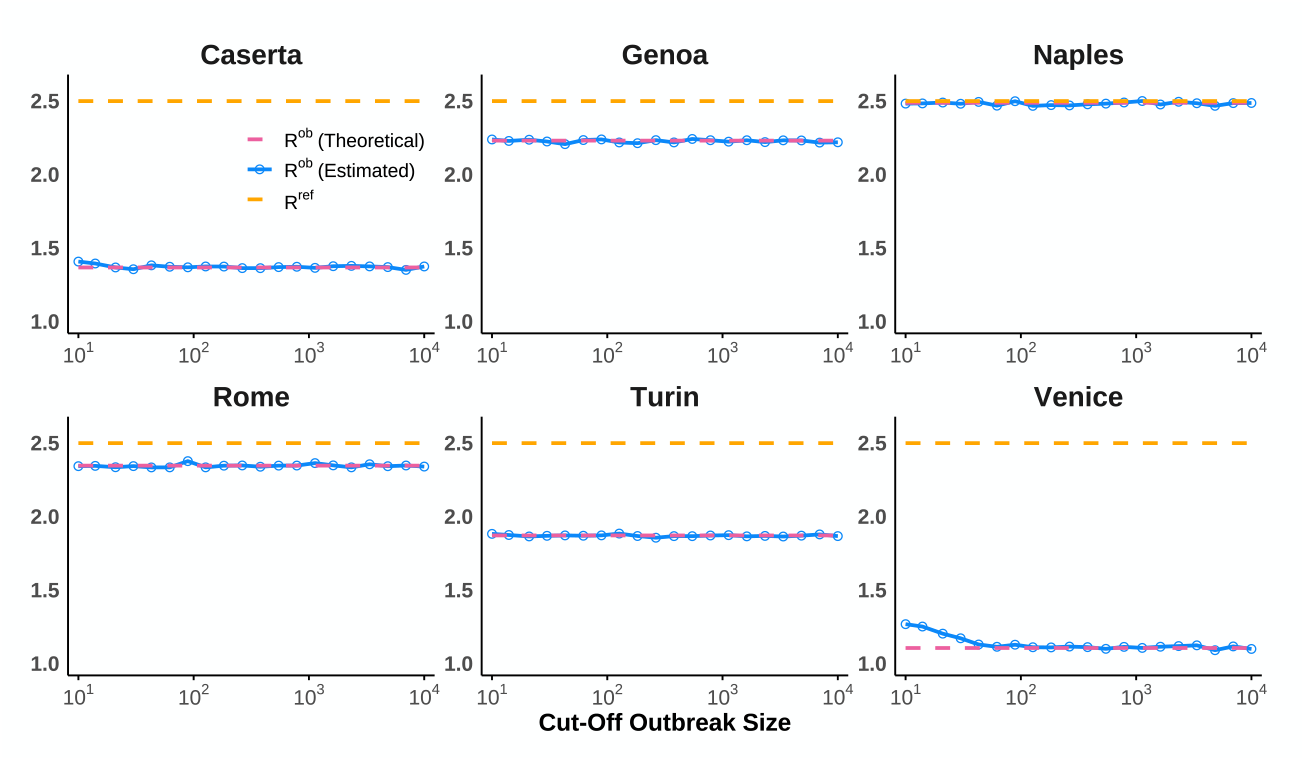
Validation of Eq.(2). The plots compare the theoretical *R*^ob^ value calculated from epidemic probabilities (Eq. (3), pink) against expectation values on outbreak data (Eq. (2), blue). Synthetic epidemic data were generated using colocation data from Italy. The x-axis represents the cut-off size used to discriminate an epidemic from extinct minor outbreak. Here, *R*^ref^ = 2.5 and expectations values were computed over 100,000 stochastic realizations.

We now show that *R*^ob^ behaves consistently as a reproduction ratio by demonstrating that it reduces to known quantities in limiting cases, and has the same critical behavior. First, the epidemic threshold of the system (*R*^ref^ = 1) correctly maps to *R*^ob^ = 1. Specifically, we prove in the Methods that, when *R*^ref^ *<* 1, then 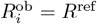 and, when *R*^ref^ *>* 1, every 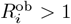. Second, from the second expression in Eq. (3), it follows directly that 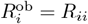 when the community is isolated, meaning that the outbreak reproduction ratio coincides with the standard local reproduction ratio. Finally, if all communities share the same total reproduction ratio (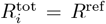 for all *i*), then 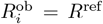 for all *i* – see the Methods for the proof. This result agrees with Ref. [15], which showed that systems lacking spatial heterogeneity in transmission potential behave effectively as a single population, whose epidemic dynamics is fully determined by *R*^ref^.

When instead transmission potential is heterogeneous, the outbreak reproduction ratio can also quantify spatial risk heterogeneity. From Eq. (3) and the Collatz-Wielandt inequalities [43], it follows that 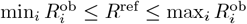. This relation shows that the outbreak reproduction ratios of individual communities are distributed around the system-level value *R*^ref^: some communities face a lower epidemic risk than predicted by global estimates, while others face a higher one.

### Estimation of the outbreak reproduction ratio from transmission chains

To estimate *R*^ob^ from surveillance data, one must account for the fact that only a single outbreak is observed, making it impossible to compute the empirical means in Eq. (2) across multiple realizations. We addressed this by first estimating the relevant entries of **R** directly from an observed transmission chain, and then computing the outbreak reproduction ratio from Eq. (3) using this estimated matrix. The detailed methodology is provided in the Methods. Figure 3 illustrates the robustness of our estimation approach using data from simulated epidemics seeded in six different Italian provinces. Expectedly, estimates were noisy and tended to overestimate *R*^ob^ on small outbreaks. They instead became more precise and tend to the corresponding theoretical values as the size of the outbreak increases. Crucially, the accuracy of the *R*^ob^ estimates depended both on the location where the outbreak originated and on the community for which *R*^ob^ was being estimated. Specifically, estimation was broadly more accurate (and requires smaller outbreaks) in major cities such as Rome and Naples. Also, accuracy was maximal if *R*^ob^ was inferred for the seeding community itself, and reliable estimates could be obtained even from relatively small outbreaks. This reflects the fact that early infections are concentrated in or near the outbreak origin. Spatial proximity, as a proxy for network connectivity, further modulated accuracy: For example, *R*^ob^ in the province of Caserta could be estimated accurately from outbreaks seeded in neighboring Naples even for relatively small outbreaks (on the order of 10^3^ infections), whereas outbreaks seeded in distant provinces such as Genoa required substantially larger outbreak sizes (exceeding 10^5^ infections), and therefore longer observation periods, to achieve comparable estimation accuracy (Fig. 3).

**Figure 3.**
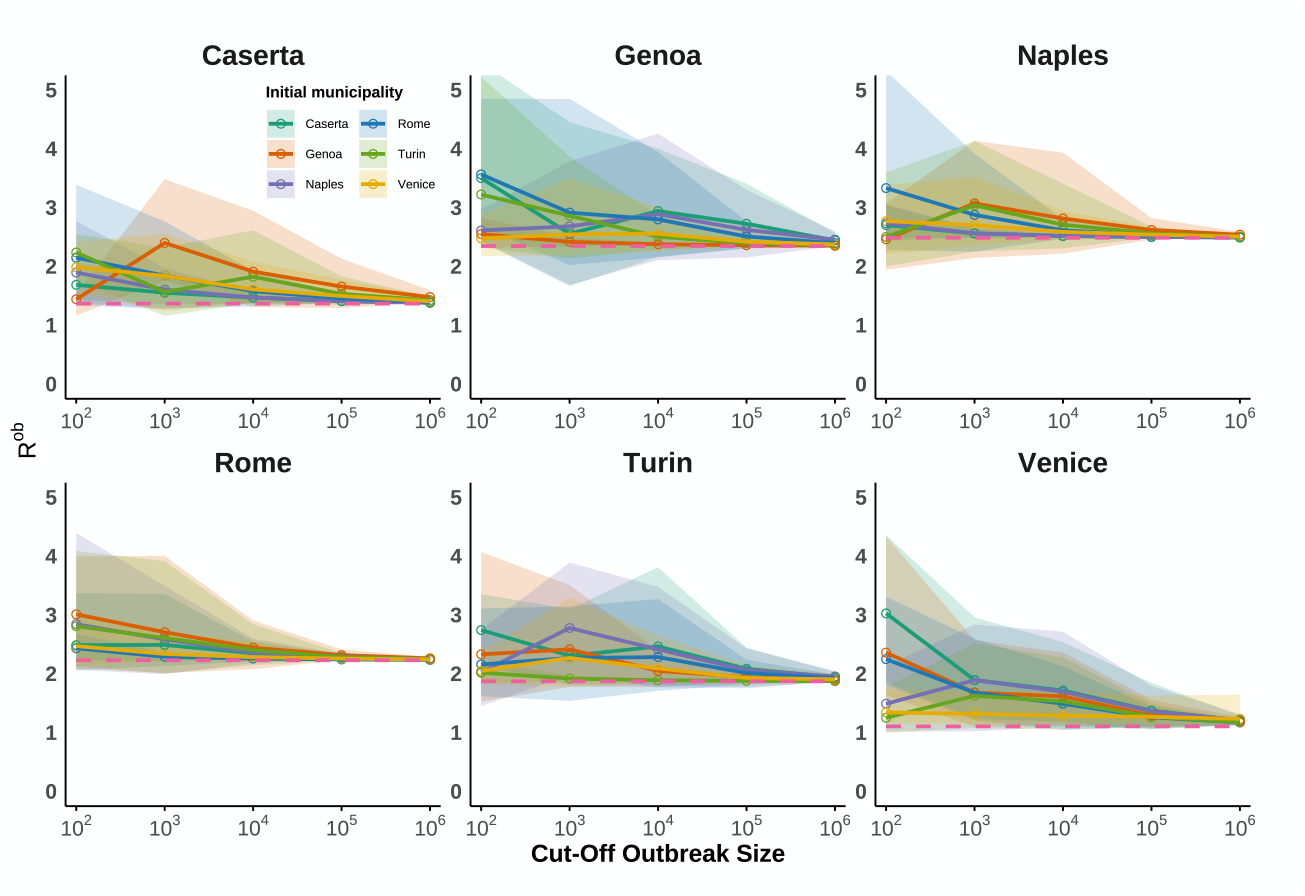
Estimation of *R*^ob^ from outbreak data. Each panel shows the estimated *R*^ob^ for a specific province as a function of the cumulative outbreak size available for analysis (x-axis), and of the seeding province (legend). Shaded regions span 90% uncertainties. Theoretical values (Eq. (3)) are represented by pink dashed lines.

These results highlight that the reliability of *R*^ob^ estimation is governed by both the size of the available dataset and the interplay between the outbreak’s origin and the underlying spatial network structure.

### Estimating outbreak risk across countries and communities using spatial contact data

Equation (3) enables the estimation of epidemic risk even before outbreaks occur, by informing **R** with data on disease characteristics and the population-level spatial contact data introduced earlier (see also the Methods). Using this approach, we estimated epidemic risk across 943 communities in 13 countries, showing that conventional metrics fail to capture the heterogeneity and distribution of epidemic risk, whereas *R*^ob^, when combined with large-scale contact data, effectively captures this heterogeneity at minimal computational cost. We explored three reference reproduction numbers: *R*^ref^ = 1.5, consistent with estimates for the 2009 H1N1 influenza pandemic strain [44]; *R*^ref^ = 2.5, corresponding to early estimates for the historical SARS-CoV-2 strain [6]; and a higher *R*^ref^ = 4, representing a hypothetical highly transmissible emerging virus. The countries included in the study appear in Fig. 4a. Figure 4b displays their gross domestic product (GDP) per capita and Fig. 4c their urbanization profile in terms of the *DEGURBA* degree of urbanisation, which classifies territories in seven categories according to a rural/urban continuum, accounting for population size and density [45].

**Figure 4.**
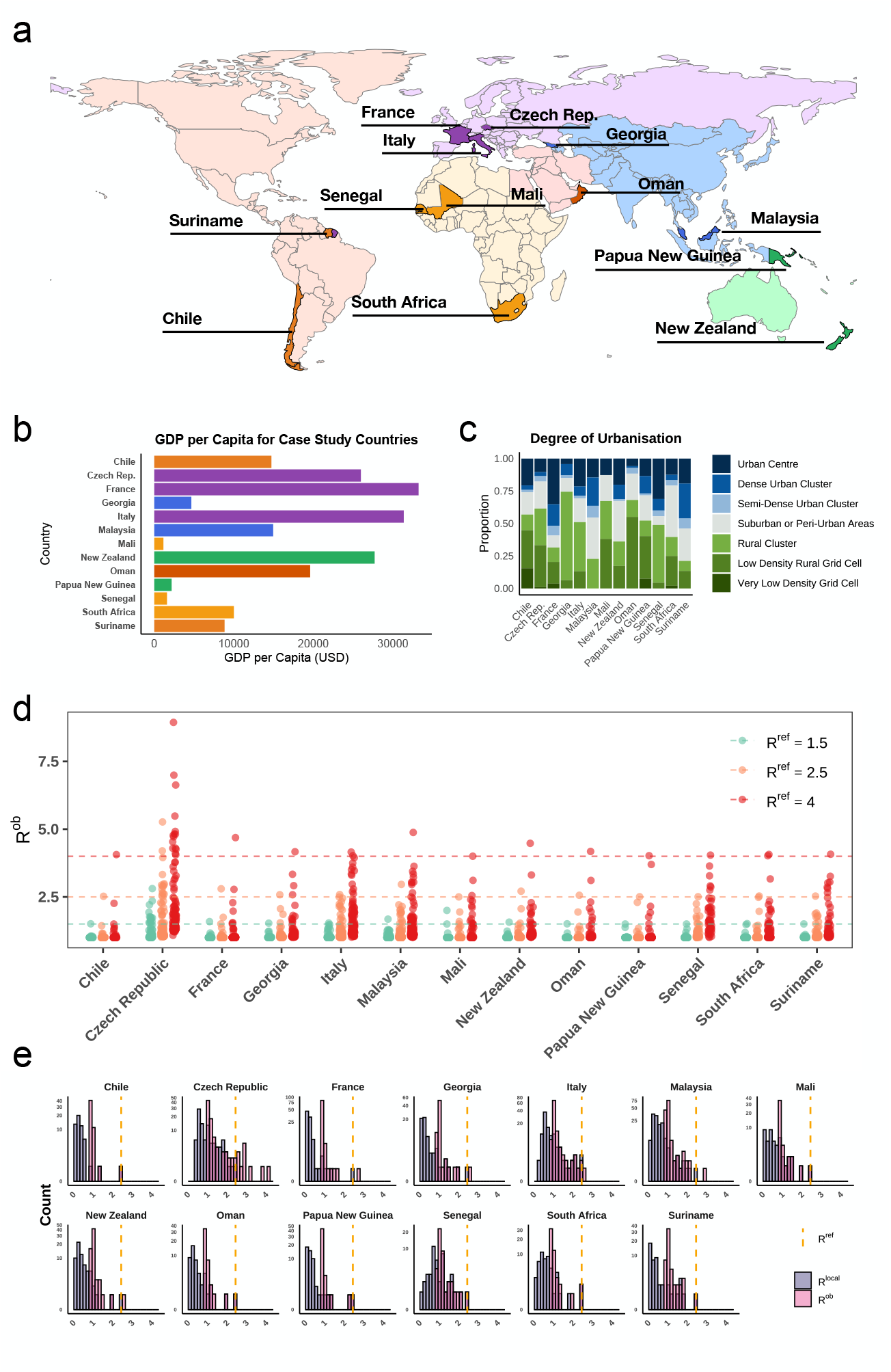
Pre-outbreak estimate of *R*^ob^ across countries, urbanization and income levels using spatial contact data. **a**. Countries included in the study, representing different WHO regions. **b**. GDP per capita of the selected countries in USD in 2022. **c**. Urbanization breakdown for each country categorized according to the Degree of Urbanisation (DEGURBA) classification of its ADMIN-2 areas. **d**. Distribution of *R*^ob^ for all ADMIN-2 spatial communities within each country under three scenarios: *R*^ref^ = 1.5, 2.5, 4.0. Each dot represents a community. **e**. Histograms comparing the distributions of the local reproduction number (*R*_*ii*_, grey bars) and *R*^ob^ (purple bars) for a scenario where *R*^ref^ = 2.5 (dashed orange line).

Values of *R*^ob^ were highly heterogeneous in all countries (Fig. 4d), indicating that country-level risk estimates are poor predictors of local epidemic potential. In several countries, epidemic risk was concentrated in a small number of communities with *R*^ob^ *> R*^ref^, while the majority of communities exhibited *R*^ob^ values close to one, implying a low probability that local emergence would trigger a large-scale outbreak. Other countries, such as Italy, displayed a broad distribution of *R*^ob^ centered around *R*^ref^, indicating heterogeneous but more evenly distributed risk. In all cases, increasing *R*^ref^ led to a widening of the *R*^ob^ distribution, with a marked increase in the maximum *R*^ob^ values.

Figure 4e demonstrates that risk measures based solely on local within-community predictors are also inadequate. Local reproduction ratio estimates (*R*_*ii*_) were consistently lower than *R*^ob^, confirming that local metrics systematically underestimate risk. Moreover, the distinct shapes of the two distributions indicate that *R*^ob^ is not merely a scalar multiple of local risk; crucially, the rank ordering of communities changes when spatial coupling is considered.

To further quantify the discrepancy between local risk estimates and *R*^ob^, we defined the local error as the relative increase in the estimated reproduction ratio obtained by using *R*^ob^ instead of a local metric. Here, local risk was measured by *R*_*ii*_, the reproduction ratio accounting only for infections generated within the same community (see Methods). In the Supplementary Information, we also evaluate *R*^tot^, which includes all secondary infections generated by local cases, as an alternative local metric, obtaining similar results ((see Supplementary Figs. 1–3)).

Figure 5a shows that local errors were systematically lower in urban areas and substantially higher in rural and low-population-density settings, across all values of the reproduction ratio considered. This indicates that in high-density environments local transmission potential is a relatively accurate proxy for epidemic risk, whereas in low-density settings epidemic risk is driven to a larger extent by spatial coupling and network-mediated transmission.

**Figure 5.**
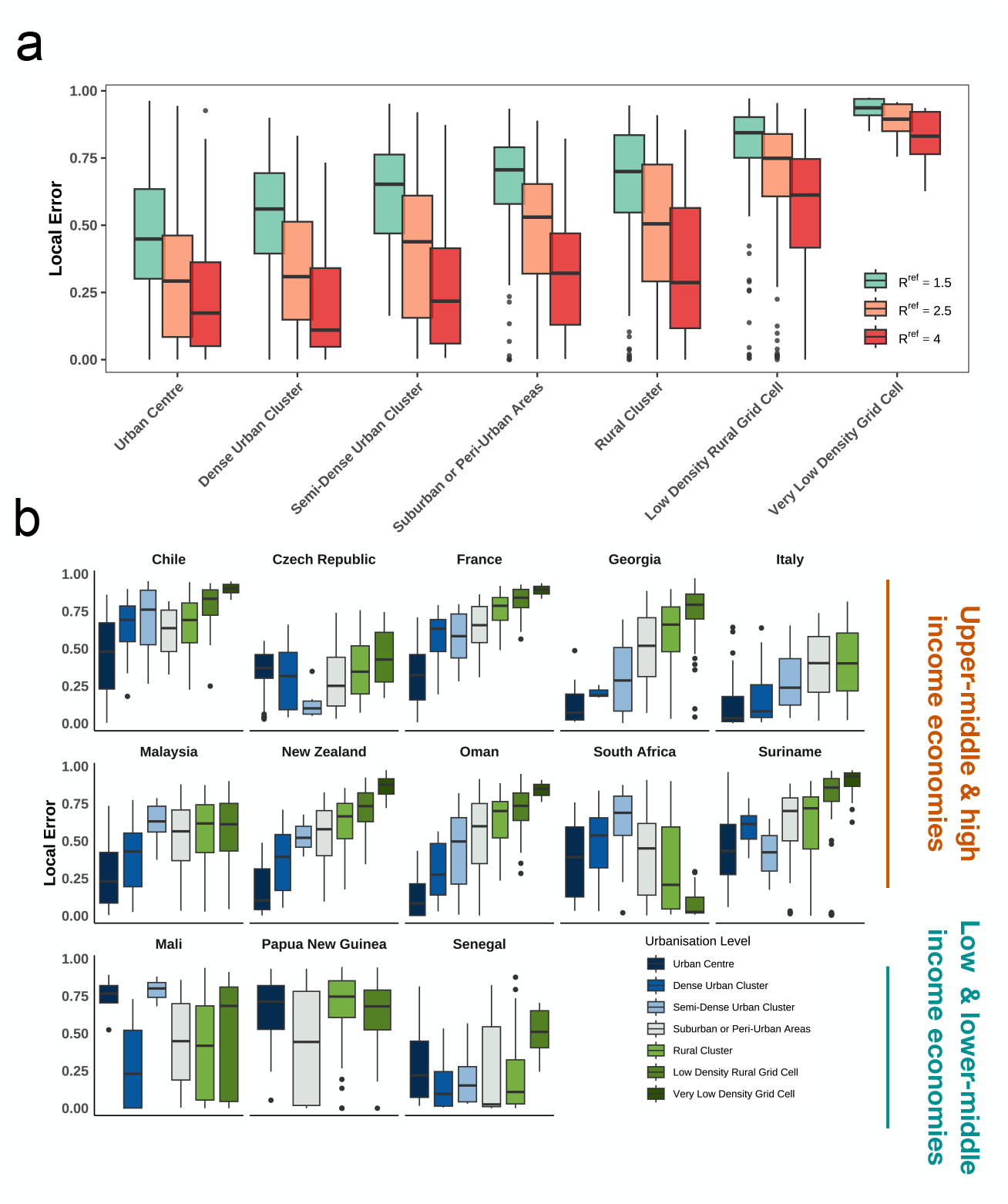
Discrepancy between local *R* and *R*^ob^ across countries. **a**. Relative local error (defined as the normalized difference between *R*^ob^ and *R*_*ii*_) stratified by urbanisation level for *R*^ref^ = 1.5, 2.5, 4.0. **b**. Local error in each country (stratified by country income group) across DEGURBA classes of urbanization.

**Figure 6.**
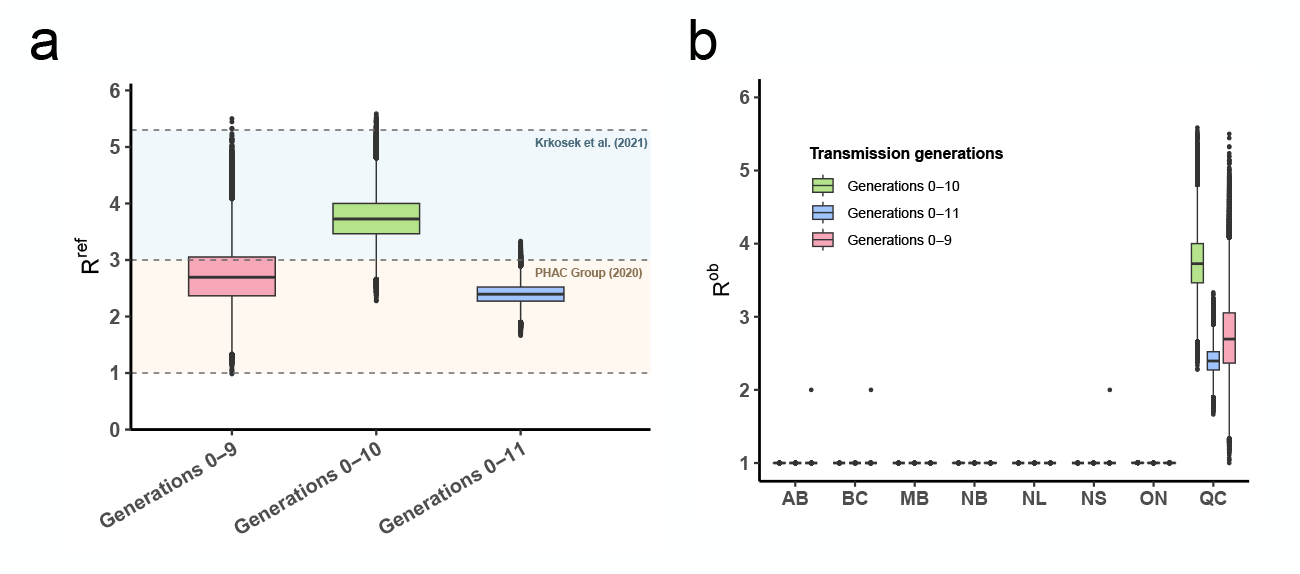
Retrospective analysis of early SARS-CoV-2 transmission in Canada. **a**. Estimates of the reference reproduction ratio (*R*^ref^) inferred from transmission trees (reconstructed from phylogenatic data), truncated at different generations 9, 10 and 11. Colored shaded regions indicate 95% uncertainty intervals of *R*^ref^ estimates from previous literature [49, 50]. **b**. Estimates of *R*^ob^ in the eight Canadian provinces for which data were available. Box plots show the 95% uncertainty posterior estimates across different data subsets. Province abbreviations: AB, Alberta; BC, British Columbia; MB, Manitoba; NB, New Brunswick; NL, Newfoundland and Labrador; NS, Nova Scotia; ON, Ontario; QC, Quebec.

There were, however, exceptions to this pattern, particularly among low- and lower-middle-income countries (Fig. 5b), where local errors remained largely uniform across urbanisation levels. This is consistent with what previously found in across African countries where mobility-driven epidemic risk was found to be substantial across diseases and transmission routes, from malaria [46], to HIV [35], to COVID-19 [47]. Among upper-middle-income countries, South Africa emerged as an outlier: local errors were comparatively low in both highly urban and highly rural settings, and instead peaked at intermediate levels of urbanisation.

### Application to early SARS-CoV-2 transmission in Canada

We estimated *R*^ob^ of early COVID-19 (March to May 2020) across Canadian provinces using available estimated SARS-CoV-2 transmission chains from phylogenetic data [48]. We included eight Canadian provinces for which data were available: Alberta, British Columbia, Manitoba, New Brunswick, Newfoundland and Labrador, Nova Scotia, Ontario, Québec. We also incorporated available colocation data as priors on the inter-community contact network to mitigate the effects of sparsity in the observed transmission chains (see Methods). Estimates of *R*^ref^ were mildly sensitive to the depth of the considered transmission chains but remained compatible with those reported in previous studies [49, 50].

Province-specific estimates of *R*^ob^ were close to the threshold value *R* = 1 in seven of the eight provinces, indicating a low probability that infections there would generate large-scale transmission. Québec was the sole exception, with *R*^ob^ consistently exceeding 2, identifying it as the primary contributor to epidemic risk in Canada. Notably, this result is consistent with retrospective reconstructions of the effective reproduction number (*R*_*t*_), which identified Québec as the only province sustaining persistent high-level transmission during the first wave of COVID-19 [49, 50].

## Discussion

Our study addressed the problem of quantifying epidemic outbreak risk in spatially structured populations before and during the early phase of spread. Epidemic risk is often synthesized through the reproduction ratio, which is easily interpretable and underpins both theoretical understanding and operational decision-making across pathogens, settings, and intervention contexts. In spatially structured systems, however, epidemic dynamics are shaped by the interplay between local transmission and mobility-mediated coupling across communities. Standard reproduction ratios, whether defined locally or at the system level, capture only partial aspects of this interplay and may therefore provide a distorted picture of outbreak risk following pathogen emergence or introduction. Specifically, local measures, such as those recently used to assess arboviral epidemic risk [9], neglect the contribution of mobility-driven transmission beyond the community of emergence, while system-level measures, such as those used for national-scale analyses of COVID-19 transmission in Italy[38], are insensitive to the initial conditions of the outbreak, despite the fact that early epidemic dynamics are strongly influenced by stochastic effects and by the location of introduction. A common way to address these limitations has been to rely on large-scale, data-driven computational models that explicitly represent contact networks and high-resolution (possibly individual-level) transmission. These approaches can reproduce realistic epidemic trajectories, but they are data intensive, require extensive parametrization, and typically move away from synthetic and interpretable risk indicators, often foregoing reproduction-ratio-based metrics altogether [40], in favor of more complex and context-specific simulation-based assessments of epidemic potential [51][39]. The results presented here overcome this limitations in the form of the outbreak reproduction ratio *R*^ob^, a parsimonious and interpretable reformulation of the reproduction ratio that integrates local transmission conditions with system-level spatial interactions.

The central result is that outbreak risk following an introduction in a given community cannot, in general, be inferred from the expected number of secondary infections generated locally or in total. Instead, it depends on where those infections occur and on the capacity of the recipient communities to sustain onward transmission. The outbreak reproduction ratio makes this dependence mathematically and operationally explicit using the framework of multitype branching processes. In this sense, *R*^ob^ is neither a purely local nor a system-level indicator: it is a community-specific measure that embeds local transmission conditions within the system-wide structure encoded in the underlying spatial contact network. This formulation clarifies why both local and global reproduction ratios may lead to systematic mischaracterizations of epidemic risk. A community with subcritical local transmission, for example, may nonetheless present a non-negligible probability of triggering a large-scale epidemic if it is connected to highly vulnerable parts of the system. The analytical properties of *R*^ob^ reflect this structure. It reduces to the standard local reproduction ratio in the absence of spatial coupling, and to the reference (global) reproduction ratio when transmission potential spatially is homogeneous, while in heterogeneous systems it naturally distributes around the system-level value, identifying communities that are structurally more or less likely to act as epidemic triggers.

The empirical analyses presented in this study illustrate the practical consequences of these properties. First, they showed that *R*^ob^ can be estimated prior to pathogen emergence or introduction using aggregated large-scale contact and mobility data. Such data, including those from Meta used here, are anonymized and widely available, enabling a privacy-preserving, lightweight, and interpretable characterization of spatially resolved epidemic risk, including in resource-constrained settings with limited data penetration and computational resources. Second, they presented a rich and heterogeneous phenomenology of epidemic risk. Across multiple countries and transmission scenarios for directly-transmitted respiratory pathogens, *R*^ob^ revealed substantial heterogeneity that neither local nor system-level metrics could capture. In particular, the discrepancy between local reproduction ratios and outbreak reproduction ratios was modulated by spatial connectivity and urbanicity, exhibiting different patterns among higher- and lower-income countries.

Our results showed that *R*^ob^ can be estimated also after introduction or emergence have occurred, from early outbreak surveillance data, although the precision of these estimates depends on both outbreak size and configuration of the spatial contact network. Analysis of simulated epidemic data showed that *R*^ob^ estimates are most reliable for the seeding community and for communities that are strongly connected to it, consistent with the concentration of early infections along dominant transmission routes. Then, the retrospective application of *R*^ob^ to real phylogenetic data describing early SARS-CoV-2 transmission in Canada illustrated how *R*^ob^ can correctly identify local epidemic potential even from during early epidemic phases.

Our findings present the outbreak reproduction ratio *R*^ob^ as a natural extension of the reproduction ratio concept to spatially structured populations. Rather than replacing existing indicators, *R*^ob^ provides a complementary quantity that explicitly encodes the contribution of spatial coupling to local outbreak risk. Its definition is independent of disease-specific natural history assumptions and relies on the reproduction operator as a unifying object, making it applicable across transmission routes and epidemiological contexts, provided that the data required to parametrize it are available [15]

This study has limitations. First, the estimation of *R*^ob^ is only as reliable as the data used to construct it. Estimates based on surveillance data are sensitive to reporting delays, underdetection, and spatially heterogeneous ascertainment, while estimates based on spatial contact data depend on assumptions linking observed proximity to effective transmission. Colocation data, in particular, provide an aggregate proxy for mixing and may suffer from representativeness bias [52]. These limitations directly propagate to *R*^ob^ and must be accounted for when interpreting quantitative values. Second, the theoretical framework does not include explicit overdispersion in secondary infections. Offspring distributions are assumed to be Poisson, with heterogeneity arising from population structure [20–22, 42, 53]. While this is enough to account for many sources of heterogeneity that are typically effectively modeled with overdispersed distributions, it does not account for individual-level variations in contact patterns and transmissibility or susceptibility profiles [25, 27, 42, 54, 55]. Third, the analysis relies on a linear approximation in which the reproduction operator is effectively constant. This approximation is appropriate for the early phase of an outbreak or for periods without major changes in immunity, behavior, or interventions, but it is not intended to describe later epidemic phases where such feedbacks substantially alter transmission [56].

In conclusion, this work introduces an analytical and interpretable framework for quantifying epidemic risk in spatially structured populations. The outbreak reproduction ratio *R*^ob^ reformulates the reproduction ratio to explicitly account for spatial coupling, and provides a lightweight, generalizable, and locally resolved yet system-aware indicator of epidemic risk that remains tied to established epidemiological concepts. This approach enables spatially resolved risk assessment using readily available data and relying on very limited computational resources, clarifying the conditions under which standard metrics fail to capture the mechanisms governing early epidemic spread, and how those limitations can be overcome.

## Data Availability

All data produced in the present study are available upon reasonable request to the authors

## Acknowledgements

Colocation data were available thanks to Data For Good at Meta.

## Funding

This study was supported by: Horizon Europe grant SIESTA (101131957) to E.V.

## Author contributions

B.W. and E.V. conceived and designed the study. E.V. developed the theory. B.W. analyzed the data, developed the code, performed the simulations. B.W. and E.V. analyzed and interpreted the results. B.W. and E.V. wrote the article.

## Methods

### Multi-Type Branching Process

Let *X*_*i*→*j*_ be the number of secondary infections that an infected resident of *i* generates in *j*. By definition of the reproduction operator, 𝔼 [*X*_*i*→*j*_] = *R*_*ji*_. Let *p*_*i*_ be the previously defined epidemic probability, i.e., the probability that a realization of the multitype branching process starting with one initial case in *i* does not go extinct. These probabilities obey the following equation (Ref. [41]):

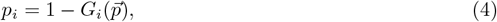

where 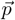 is the vector of all *p*_*i*_s, and *G*_*i*_ is the multivalued probability-generating function of the stochastic vector 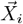, encoding the number of secondary infections from a resident of *i* to every community: 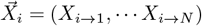.

Let us assume that *X*_*i*→*j*_ are Poisson-distributed and that *X*_*i*→*j*_, *X*_*k*→*l*_ are independent unless *i* = *j* and *k* = *l*. Recalling that the probability generating function of the Poisson distribution with mean *μ* is *g*(*z* | *μ*) = *e*^*μ*(*z*−1)^, we can compute the probability-generating function in Eq. (4): 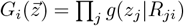. Substituting in Eq. (4), one gets Eq. (1).

### Spatial contact data to model the reproduction operator

We estimated the reproduction operator **R** using Colocation Maps provided by Meta Data For Good [52]. Colocation Maps use the probability, computed from mobile device usage, that a randomly chosen resident of community *i* and a randomly chosen resident of community *j* find themselves in the same 600 m *×* 600 m tile during a randomly chosen five-minute time window. The data were provided at the ADM 2 level and for week 13 of 2023. We combined Colocation Maps with population data and built the reproduction operator (up to a constant), following what we had done in Ref. [15]. Specifically, with *C*_*ij*_ being the colocation rate provided by Meta and *n*_*i*_ the population of spatial communities, then *R*_*ij*_ ∝ *C*_*ij*_*n*_*i*_. The arbitrary multiplication constant was then used to set the reference reproduction ratio *R*^ref^, i.e., the spectral radius of **R**.

### *R*^ob^ as a function of the epidemic probabilities

Our first goal is to compute 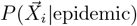, which will then be used to compute the expectation values in Eq. (2). We use Bayes’ theorem:

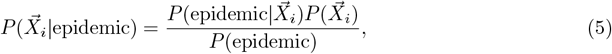

and then compute each term:

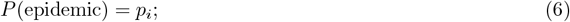

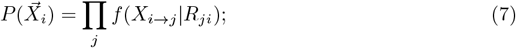

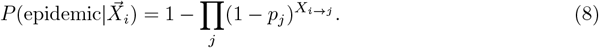

Here, *f* (*X*_*i*→*j*_|*R*_*ji*_) is the probability mass function of a Poisson distribution with mean *R*_*ji*_. The term 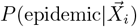 is the probability that at least one of the secondary infections triggers a large-scale epidemic. This gives

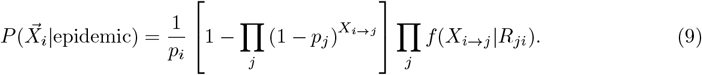

We use this to compute the expectation value:

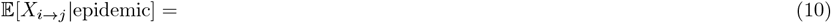

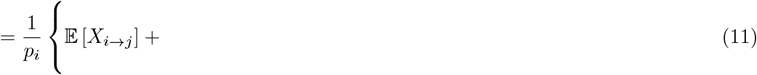

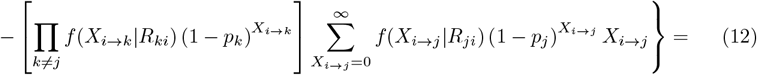

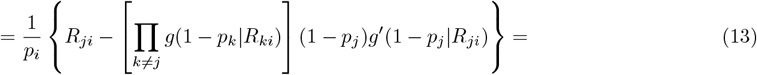

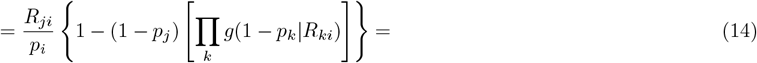

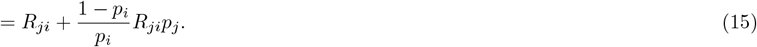

Here, we have used that Σ _*X*_*f* (*X* | *r*)*Xz*^*X*^ = *zg*^*/*^(*z* | *r*), with *g* being the probability-generating function of *f* . Also, given that *f* is Poisson and *g*(*z*|*r*) = *e*^*r*(*z*−1)^, Σ _*X*_ *f* (*X*|*r*)*Xz*^*X*^ = *zrg*(*z*|*r*). We have also used Eq. (1) in the last line.

From the identity

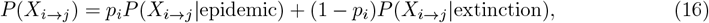

combined with Eq. (15), one can derive

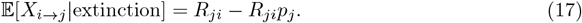

The difference between Eq. (15) and Eq. (17), summed over *j*, proves the second expression in Eq. (2). The first expression is then also recovered simply by replacing Σ_*j*_ *R*_*ji*_*p*_*j*_ = − log(1 − |*p*_*i*_), which comes from Eq. (1).

### Homogeneous transmission potential

We wish to prove that the total reproduction ratio of each community is the same 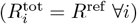 if and only if 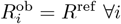.

First we prove that 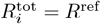 if and only if *p*_*i*_ = *p*, for *p* solving 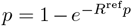. If the former is true then **R** is left-stochastic (up to a scalar) and a vector of ones is the Perron eigenvector of **R**^*T*^, and *R*^ref^ is the Perron eigenvalue. This means that Eq. (1) collapses to 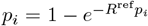 for all *i*, and *p*_*i*_ = *p*. If the latter is true then Eq. (1) implies that Σ_*j*_ *R*_*ji*_*p*_*j*_ = *pR*_*ji*_, implying again that a vector of ones is a positive eigenvector of **R**^*T*^ . This means that the columns of **R** sum to the same quantity, which must be *R*^ref^ because that eigenvector must be the Perron eigenvector.

Now we prove that if 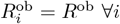 then *R*^ob^ = *R*^ref^. If the hypothesis is true, Eq. (3) implies that Σ_*j*_ *R*_*ji*_*p*_*j*_ = *R*^ob^*p*_*j*_. Then *R*^ob^ is the Perron eigenvalue of **R**^*T*^ and **p** the Perron eigenvector. Using this in Eq. (1) one gets that *p*_*i*_ = *p*. Then, each column of **R** sums to *R*^ref^, which is also the Perron eigenvalue. Then, *R*^ob^ = *R*^ref^.

Finally, we prove that if the columns of **R** all sum to *R*^ref^ then 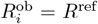. If the hypothesis is true then *p*_*i*_ = *p* and again *R*^ob^ must be equal to the Perron eigenvalue, by virtue of Eq. (3).

### Continuation of the outbreak reproduction ratio below the system’s epidemic threshold

Let *p*_*i*_(*t*) the probability that the epidemic has not died out by generation *t*. It obeys the following iterative equation:

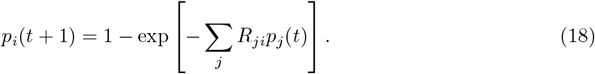

Its stable, fixed-point solution defines the epidemic probability as in Eq. (1): *p*_*i*_ = lim_*t*→∞_ *p*_*i*_(*t*) [41]. Also, let us assume **R** is nonnegative and irreducible, i.e., it is interpretable as the adjacency matrix of a strongly connected directed graph, so that Perron-Frobenius theory applies. If this were not the case the analysis could be performed separately on each strongly connected component [15]. Below the epidemic threshold (*R*^ref^ *<* 1), *p*_*i*_ = 0 ∀*i*, preventing from finding *R*^ob^ directly from Eq. (3) in that regime. We will use the time evolution to perform a continuation of the outbreak reproduction ratio to below the epidemic threshold. Eq. (3) can be trivially written as follows:

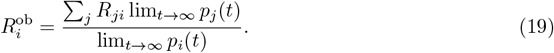

The continuation consists in replacing this with the following equation, swapping the time limit and the computation of the outbreak reproduction ratio:

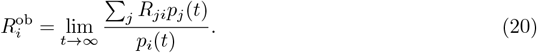

When *R*^ref^ *>* 1 and *p*_*i*_ *>* 0 the two expressions are trivially identical and we can use the latter to go to *R*^ref^ *<* 1. If *t* is large enough, *p*_*i*_(*t*) are small so that we can linearize Eq. (18) around the fixed-point solution *p*_*i*_ = 0 (we are *R*^ref^ *<* 1) keeping 𝒪 (*p*_*i*_(*t*)):

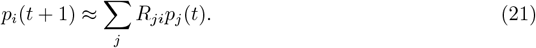

This means that

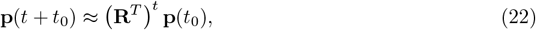

for some *t*_0_. Let us now define **v** as the right Perron eigenvector of **R** and **v**^*^ is dual vector (left Perron eigenvector), normalized so that **v**^*^**v** = 1. Then, if *t* is large enough, Perron-Frobenius theory states that (**R**^*T*^) ^*t*^ ≈ (*R*^ref^)^*t*^**v**^*T*^ **v**^*,*T*^, with **a, b** right and left Perron eigenvectors of **R**^*T*^ and **b**^*T*^ **a** = 1. Inserting into Eq. (22), one gets

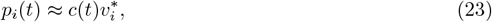

where *c*(*t*) is some scalar with *c*(∞) = 0. This relegates the time dependence to a scalar prefactor, and the limit in Eq. (20) can now be performed.

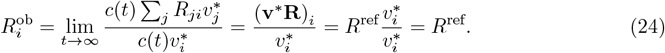

### Stochastic, spatially explicit epidemic model

To validate the equivalence between Eq. (2) (the definition of the outbreak reproduction ratio) and Eq. (3) (its expression in terms of epidemic probability), we simulated the early phase of an epidemic by sampling from the multitype branching process. We used Colocation Maps from Italy to inform **R**, as previously explained.

First, we computed *p*_*i*_ by numerically solving Eq. (1) and then computed *R*^ob^ from Eq. (3). For each spatial community *i*, we ran 100, 000 simulations started with one infected individual in *i*. We ran each simulation to extinction or upon reaching a cutoff epidemic size *M* . Simulation outcomes were classified as either *epidemic*, if the outbreak size exceeded the cutoff, or *extinction*, if it did not. From those, we computed 𝔼 [ Σ_*k*_ *X*_*i*→*k*_|epidemic] and 𝔼 [Σ _*k*_ *X*_*i*→*k*_|extinction] as empirical means and estimated *R*^ob^ from Eq. (2).

### Estimating the outbreak reproduction ratio from outbreak data

To estimate *R*^ob^ using data from a single outbreak, we first infer **R** from *T* + 1 generations of the branching process (*t* = 0, … *T* ). Let *Y*_*i*→*j*_(*t*) be the observed number of infections in j at time step *t* generated by residents of *i*, and let *I*_*i*_(*t*) = Σ _*j*_ *Y*_*j*→*i*_(*t*) be the number of incident infections in *I* at time step *t*. We know that *Y*_*i*→*j*_(*t*) is Poisson-distributed with mean equal to *I*_*i*_(*t* − 1)*R*_*ji*_, and that generation along different links is independent. This gives the following likelihood:

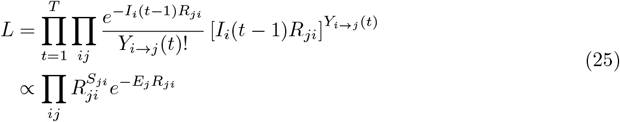

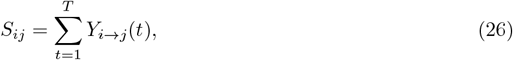

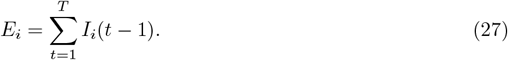

where we introduced the sufficient statistics *S*_*ij*_, *E*_*j*_. The likelihood thus factorizes into a product of Gamma-distributed *R*_*ij*_ with shape *α*_*ij*_ = *S*_*ji*_ − 1 and rate *β*_*ij*_ = *E*_*j*_, leading to the maximum-likelihood estimator of *R*_*ij*_ (see also Ref. [57]):

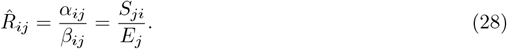

Then, by sampling from the Gamma distributions of the individual *R*_*ij*_ entries and computing *R*^ob^ through Eq. (1) and Eq. (3) for each sample, we reconstructed the empirical distribution of the outbreak reproduction ratios.

In addition, it is interesting to study how the uncertainty on *R*_*ij*_ evolves with the available data:

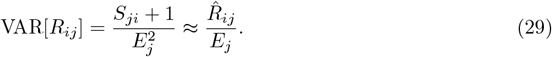

At large enough *T*, 𝔼[*I*_*i*_(*t*)] ≈ [**v**^*^**I**(0)](*R*^ref^)^*t*^*v*_*i*_. Assuming that the initial infection is in community *i*_0_, this means 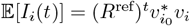. This implies that, at large *T*,

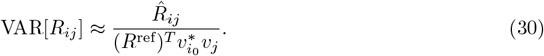

This implies that the precision at which we can estimate *R*_*ij*_ i) increases with the depth of the transmission chain that we observe, ii) is higher in the communities where the asymptotic equilibrium distribution of infections (see Ref. [15]) is concentrated, iii) is higher if seeding occurs where the dual equilibrium distribution is concentrated.

### Geography of risk estimates

Our analysis included Chile, the Czech Republic, France, Georgia, Italy, Mali, Malaysia, New Zealand, Oman, Papua New Guinea, Senegal, Suriname, and South Africa, spanning different continents and income levels.

To identify structural factors driving discrepancies between *R*^ob^ and naive risk estimates, we considered the following relative discrepancies:

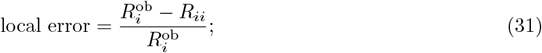

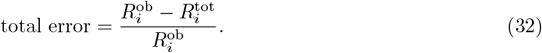

The analysis with *local error* is reported in the main text, the analysis with *total error* in the Supplementary Information.

### Inference from transmission trees in Canada

We analyzed transmission trees inferred from SARS-CoV-2 genomic sequences collected in Canada between March and May 2020, covering generations 0 to 11 of transmission [48]. To link these trees (where communities are defined at the provincial level) to the spatially explicit framework developed above, we adopted a hybrid inference scheme in which the spatial structure is fixed by contact data and only the overall transmission intensity is inferred.

We used the normalized colocation matrix **M** derived from Meta Colocation Maps, with entries *M*_*ij*_ ∝ *C*_*ij*_*n*_*i*_ and spectral radius *ρ*(**M**) = 1 (see Spatial contact data in the Methods). We assumed the reproduction operator takes the form **R** = *R*^ref^**M**, where *R*^ref^ is a scalar reference reproduction ratio. Under this parametrization, the Poisson likelihood in Eq. (25) simplifies to a one-parameter model in *R*^ref^, utilizing the sufficient statistics *S*_*ij*_ and *E*_*i*_ derived from the transmission trees (see Eqs. (26)–(27)). We assigned a flat prior to *R*^ref^ (uniform over positive real values) and sampled from the posterior distribution using Markov Chain Monte Carlo in Stan [58]. For each posterior draw, we reconstructed **R**, solved Eq. (1) for the epidemic probabilities *p*_*i*_, and computed *R*^ob^ via Eq. (3). The procedure was repeated after truncating the trees at generations 0-9 and 0-10 to assess the robustness of estimates to chain depth.

## Notes

### Competing Interest Statement

The authors have declared no competing interest.

### Summary of Updates

New version on figures mainly on figure5.

